# Contributions to the force of infection of SARS-CoV-2 in Dutch long-term care facilities

**DOI:** 10.1101/2024.11.19.24316339

**Authors:** Mariken M. de Wit, Marino van Zelst, Tjarda M. Boere, Rolina D. van Gaalen, Mart C. M. de Jong, Albert Jan van Hoek, Quirine ten Bosch

## Abstract

**Background:** Residents of long-term care facilities (LTCFs) have been disproportionately affected during the COVID-19 pandemic. To inform decision-making around interventions, we quantified the SARS-CoV-2 infection risk for residents and the relative contribution of different infection sources. We estimated the force of infection (FOI) experienced by Dutch LTCF residents over time and quantified the contribution of residents, LTCF healthcare workers (HCWs), and the general population.

**Methods & findings:** Case data were obtained by Municipal Health Services as part of the Dutch national surveillance program. During the study period (1 October 2020 to 10 November 2021), testing policies included symptom-based testing, exposure-based testing, and facility-wide serial testing. We used a data augmentation approach to include uncertainty in the timing of infection, while taking account of different testing policies. We constructed a Bayesian generalized linear model to estimate group-specific transmission rate parameters and contributions to the FOI experienced by residents.

During the study period 36,877 cases were registered among residents and 19,676 among HCWs. The total daily FOI towards residents was highest in December 2020 (1.7*10^−3^, 95% CI: 1.5*10^−3^ – 1.9*10^−3^) and lowest in June 2021 (1.1*10^−5^ 95%CI: 7.6*10^−6^ – 1.7*10^−5^). Resident-directed type-reproduction numbers and FOI declined as COVID-19 vaccination rollout started in residents, HCWs, and the older general population (February-May 2021). Most resident infections in spring and summer 2021 were attributable to infections in the general population. The relative contribution of the general population to the FOI decreased in summer 2021 when vaccination was available population-wide. In October-November 2021, type-reproduction numbers and FOI increased again. We observed an increase in residents’ susceptibility to infection in this period, which was only partially explained by the emergence of the Delta variant. Sensitivity analyses showed that the temporal trends in relative contributions to the FOI were not impacted by assumptions about immunity build-up among residents, nor by underreporting of infections.

**Conclusions:** COVID-19 vaccination appears to have been effective in reducing SARS-CoV-2 transmission towards residents, although other factors such as seasonality or non-pharmaceutical interventions may also have contributed to this. This effect seemed to have decreased by autumn 2021, which could be due to waning of immunity or changes in control practices. Our estimates of temporal trends in relative contributions to the FOI in LTCF residents can help target intervention efforts.

## Introduction

Long-term care facilities (LTCFs), which house amongst the most vulnerable people in our societies, have been disproportionately affected during the Coronavirus disease (COVID-19) pandemic. LTCF residents are faced with increased risk of high COVID-19 impact, due to a combination of risk factors for respiratory infections, such as frailty, comorbidities, crowded living conditions, and high-frequency contact with health care workers (HCWs) [1, 2]. The lack of appropriate protective equipment for HCWs in the early stages of the COVID-19 pandemic further contributed to this. The impact is reflected in high incidence and disease burden, peaking in the high age groups [3]. Seroprevalence estimates among healthcare workers (HCWs) in Dutch LTCFs also indicate high Severe Acute Respiratory Syndrome Coronavirus-2 (SARS-CoV-2) infection rates relative to the total HCW population (16.9% vs. 7.5%, after the first wave) [4]. In the first half of 2020, deaths among LTCF residents accounted for 37-66% of all COVID-19 related deaths across several EU/EEA countries [5]. Estimations of the infection fatality rate for LTCF residents in this period are high, with estimates around 22% [6].

In response to the many outbreaks in LTCFs, the WHO published a policy brief with recommendations for managing SARS-CoV-2 transmission in LTCFs [7]. Governments introduced non-pharmaceutical interventions (NPIs) such as visitor bans, individual movement restrictions, quarantine policies, masking policies, and a plethora of testing strategies [8]. However, NPIs might also affect physical and mental well-being of residents and their families and therefore require a careful balance and judicious implementation [9].

NPIs can be categorized into two groups: those focused on reducing the introduction of infections into the LTCF and those focused on reducing onwards transmission within the LTCF after introduction. The potential impact of a specific NPI depends on the contribution of introductions relative to within-LTCF transmission to the overall case load. Here, we consider within-LTCF transmission as the subsequent infections occurring after an introduction, between and among HCWs and residents. LTCFs are generally open for people to enter and leave, albeit sometimes with restrictions such as visiting hours or locked units where residents cannot exit freely for their own safety. SARS-CoV-2 outbreaks in LTCFs were found to be correlated to transmission patterns of SARS-CoV-2 in the general population [10–13]. Also, the number of HCWs at the facility, after controlling for LTCF size, was identified as a risk factor for infections among residents [14], and staff and residents were shown to be frequently infected with genetically similar SARS-CoV-2 viruses [15], indicating transmission between HCWs and residents. Similarly, the positive association between multibedded rooms as well as high occupancy rates with infection risk indicates a role for resident-to-resident transmission [16]. Such positive association has also been observed for other respiratory infections [17]. Despite evidence of transmission originating from these three groups (i.e., general population, HCWs, and residents), a quantitative understanding of their contribution to transmission and the role of interventions therein is lacking.

In this study, we aimed to disentangle the contribution of residents, HCWs, and the general population to the force of infection (FOI) experienced by LTCF residents (i.e., daily rate at which susceptible LTCF residents get infected). First, we described the characteristics of the COVID-19 epidemic in LTCFs in the Netherlands. We then used a Bayesian modelling framework including data augmentation to estimate the transmission rate parameters of each group towards residents in the period between October 2020 and November 2021. We also explored the changes in these rates over time and discuss how these relate to changes in the epidemiological context.

## Data and methods

### Setting

A total of 3.2 million SARS-CoV-2 positive tests were recorded in the Netherlands in 2020 and 2021 [18], with an estimated infection seroprevalence of 26% in November 2021 [19]. The first detection of SARS-CoV-2 infection in the Netherlands was on 27 February 2020 [20]. COVID-19 vaccine rollout started in HCW and LTCF residents ahead of the general population. Going forward, we will distinguish five periods based on progress in vaccination campaigns. The chosen dates do not represent exact starting dates of vaccination, but approximate the moment when immunity started to develop in the relevant population because of vaccination. Period A (1 October 2020 – 25 January 2021): pre-vaccination period. Period B (26 January 2021 – 28 February 2021) and period C (1 March 2021 – 30 May 2021): the periods after the first (B) and second (C) vaccination round for the primary series in LTCF residents and HCW. Period D (31 May 2021 – 30 September 2021): general population was getting vaccinated. Period E (1 October 2021 – 10 November 2021): period after vaccination of the general population. Data from before October 2020 was excluded due to limited accessibility to testing.

Dutch LTCFs for older people with high-level care needs can range from small-scaled homelike nursing homes to large residential care centers that have both nursing home and care home/service flat facilities. Typically, LTCFs have common areas for meals and socializing, and the rooms are predominantly one-bedded. LTCFs offer specialized wards, mostly either somatic wards for physically disabled residents, psychogeriatric wards for residents with dementia, or geriatric rehabilitation wards. In addition to society-wide control measures, such as physical distancing, hygiene measures, and self-quarantine, specific measures were implemented in LTCFs [21]. These included isolation and quarantine measures and visitor restrictions. Testing policies during the study period (1 October 2020-10 November 2021) included symptom-based testing, exposure-based testing, and facility-wide serial testing, although adherence to these measures varied [22].

### Data collection

Data were obtained as part of the national surveillance program, through contact tracing by the Municipal Health Service (GGD: Gemeentelijke Gezondheidsdienst). Data from 24 out of 25 GGD regions were included. We used two types of data: 1) cases in the general population were obtained from aggregated data with total number of cases (i.e., positive RT-PCR test) per GGD region per day and 2) cases in HCW and residents were obtained from an individual-level dataset with data about cases associated with LTCFs for elderly care. A case was classified as associated with an LTCF when this person appeared in an LTCF-associated cluster (i.e., most likely setting of infection based on contact-tracing information). The individual-level dataset included: sex, age, GGD region (using an anonymized ID), symptom onset date, date of positive test result, date of notification, all-cause mortality (yes/no and date), whether they lived or worked in an LTCF, and a cluster identifier (i.e., pseudonymized LTCF identifier if the LTCF was the most likely setting of infection). A large LTCF organization with multiple facility locations was viewed as consisting of multiple LTCFs, such that an LTCF refers to a specific LTCF location in this study.

Additional data sources included the age distribution of the general population (Statistics Netherlands (CBS) [23]) and the bed capacity of LTCFs obtained through Patiëntenfederatie Nederland. This information was available for 26% of LTCF locations and was linked to LTCFs by matching postal codes before further pseudonymization of the study dataset.

Resident and HCW cases associated with LTCFs (N=56,553) were included in descriptive analyses. Individuals who were recorded both as resident and HCW (n=137) were classified as resident.

### Statistical model

We aimed to quantify the force of infection (FOI) experienced by LTCF residents. The FOI (*λ*) denotes the rate at which susceptible individuals become infected per day. Assuming infections follow a Poisson process, the probability of a susceptible individual to acquire infection during a period *Δt* is related to *λ*

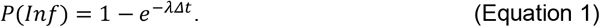

Here, *λ* depends on the fraction of individuals who are infectious and on the transmission rate parameter *β*: the average number of daily new infections per infectious individual, such that 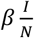, where *β* can be considered the product of the contact rate and the probability of a contact between an infectious and susceptible individual to result in infection. The latter is related to infectivity and susceptibility of the infectee and recipient, respectively.

We distinguished the contribution to *λ* by three groups of infectious individuals in LTCF: i) residents, ii) HCWs, and iii) general population. HCW were classified as such when they self-identified as HCW in an LTCF during contact tracing. The general population coming into contact with residents consisted of visitors, non-healthcare LTCF staff, and possibly HCWs in case of missing information regarding profession. We assumed *β* to differ between these groups because of differences in contact rates and infection probability upon a contact between an infectious and susceptible individual. The daily probability for a resident to become infected by either of three groups is

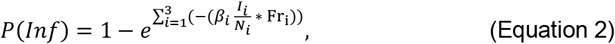

where *β*_*i*_(*t*) is the transmission rate parameter for group *i, I*_*i*_ is the number of infectious people for group *i*, and *N*_*i*_ is the population size for group *i*. Fr_*i*_ represents the fraction of overall resident contacts that are with individuals of group *i*. It was not possible to distinguish changes in the transmission rate parameter from changes in contact patterns, due to these parameters being linearly related to each other. We therefore set *Fr*_*i*_ to be 1/3 for each group, implying that deviations from this fraction will be reflected in the transmission rate parameters. We allowed group-specific transmission rate parameters to differ between periods (see Supplementary Material).

Applying equation 2 to all LTCFs in the data (*N)* and for all time points *t*, gives the likelihood function with *z* representing the data and θ representing the parameters:

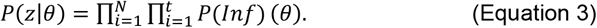

### Inference

To estimate the transmission rate parameters by group and over time, we fitted a generalized linear model to the data in a Bayesian framework. To this end, we linearized equation 2 using a complementary log-log transformation. Models with and without interaction terms between group and period were compared using the Widely Applicable Information Criterion (WAIC)[24] (see supplementary figure 4). The following equation represents the full model, including all interaction terms:

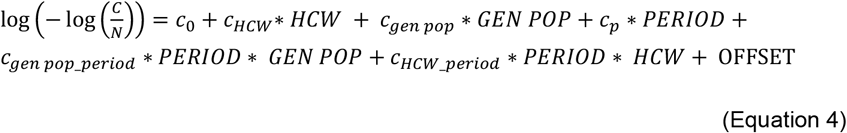

with *c* denoting the regression coefficients. Here, c_0_ denotes the intercept, with residents (*res*) and period A being the reference categories. Other coefficients relate to HCW *(hcw)*, the general population (*gen pop*), period *(p)*, and the interactions of period with the general population (*gen pop_period*) and with HCW (*HCW_period)*. The mean prevalence, 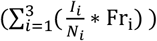, denotes the offset variable [25]. The capitalized explanatory variables for each group consist of 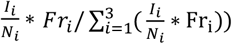, with *i* denoting the group and residents in period A acting as the reference group. The dependent variable represents the number of cases among residents (C) divided by the total resident population (N).

The model uses the daily prevalences of infectious individuals per group 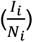as the offset variable. The daily prevalence estimates for residents and HCW were calculated at LTCF-level, whereas the daily prevalence in the general population was calculated at regional level and applied to all LTCFs within that region. The data described the timing of symptom onset or diagnostic test result, not when infections occurred and how long they persist, which is needed to calculate prevalence. We used data augmentation techniques [26, 27] to account for uncertainty in the dates of infection within the inference framework thereby creating 100 different datasets. For the aggregated number of cases detected in the general population, no data augmentation was performed, but rather we imputed the estimated day of infection. Lastly, for those cases with missing information on LTCF identifier we imputed this based on the LTCF associated with the cluster they were part of. For those clusters that were associated with multiple LTCFs, the LTCF was randomly selected amongst these for each of the datasets to account for this uncertainty. Further details on the applied augmentation techniques are described in Supplementary Material Methods.

For each of 100 augmented datasets, we fitted equation 5 in a Bayesian GLM framework with a Hamiltonian Monte Carlo sampling algorithm of 2000 iterations, discarding the first 1000 as burn-in period. Uniform priors across any real number were used for the regression coefficients. A gamma distributed prior (alpha = 0.01, beta = 0.01) was used for the overdispersion parameter and a beta distributed prior (alpha = 1, beta = 1) for the zero-inflation parameter. We calculated WAIC values [24] for each model run (i.e., each dataset) and averaged these to assess model fit. WAIC values were used to compare model variants assuming binomial, beta-binomial, zero-inflated binomial, and beta-binomial zero-inflated distributions and provide insight into the extent to which superspreading events played a role in LTCFs (i.e., resulting in overdispersion and/or zero-inflation in the data). Further analyses were done on pooled results from all model runs. All analyses were conducted in R version 4.2.1., model fitting was done using the *brms* package [28].

Period-specific transmission rate parameters from each group towards residents (*β*_*i*_(*t*)) can be derived from the regression coefficients. For example, the transmission rate parameter from the general population towards residents in period C is calculated as 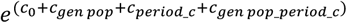. These regression coefficients can also be used to estimate the susceptibility of residents during a specific period relative to period A and the infectivity of HCW and the general population relative to residents. Relative resident susceptibility for period *i* was estimated as 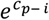. Relative infectivity of the general population was estimated as 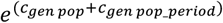 and similarly for HCW: 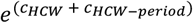. In this analysis, we corrected for an increased transmission success of the Alpha (1.7 times higher [29]) and Delta (2 times higher [30]) variants, relative to the circulating variant (Supplementary Material Methods). By doing so, we aimed to disentangle temporal changes in susceptibility due to new variants from external factors such as natural immunity and vaccination. Uncorrected results are presented in supplementary figure 10.

We used the estimated *β*_*i*_(*t*) to calculate the effective type-reproduction numbers (R) for transmission towards residents. These partial Rs indicate the average number of new infections among residents arising from one infectious individual of group *i* over the course of its infectious period. R was calculated as the product of *β*_*i*_(*t*) and the infectious period used in the analysis (5 days).

### Sensitivity analysis

Estimating the number of new cases using the probability of infection requires information on the number of susceptible residents for each day at each LTCF. The number of susceptible residents was defined as the LTCF capacity minus the number of infectious residents. We thus do not explicitly correct for the build-up of immunity. Rather, the build-up of immunity is reflected in the estimates of *β*_*i*_(*t*): the probability for an infectious contact to result in infection (*p*) is assumed to be lower if the average susceptibility to infection in a LTCF decreases, e.g. because of natural or vaccine-induced immunity. We explored the impact of the assumption regarding the size of the susceptible resident population in a sensitivity analysis. For this, we modelled immunity after infection as perfect and lasting, thereby reducing the estimated susceptible population over time. In this analysis, vaccine-induced immunity, and changes over time therein, remains reflected in *β*_*i*_(*t*) estimates Additionally, we conducted a sensitivity analysis to assess the impact of underreporting of infections in the general population.

## Results

### Descriptive analyses: LTCF-associated cases

Over the course of the study period (01-10-2020 to 10-11-2021) a total of 36,877 SARS-CoV-2 cases were registered among LTCF residents and 19,676 among HCWs (Figure 1A & C). These cases were distributed over 1787 LTCF locations in the Netherlands. The median age of residents was 86 years (interquartile range (IQR): 80-91 years) and the median age of HCWs was 46 years (IQR range: 30-56 years). Both the resident and HCW populations consisted mainly of women, with 91% of HCW and 67% of residents. Of the 5791 recorded deaths in LTCF-associated cases, 6 (0.1%) were among HCWs. Survival of infected residents differed between periods, with the highest survival in periods C and D (Figure 1G). The number of cases among residents and HCWs was strongly correlated (R^2^=0.80) (Figure 1B). This correlation was lowest in period E (R^2^=0.17). With on average 2,295 cases per week, case numbers were highest in period A. Cases among LTCF residents were weakly correlated to the number of detected infections in the general population (R2=0.27) (Figure 1D). This correlation was strongest in period E (R^2^=0.83) and weakest in period D (R^2^= -0.01). When comparing a lead and a lag of one, two and three weeks, we found that a one week lead of LTCF cases compared to the general population showed the strongest correlation (R^2^=0.32).

**Figure 1:**
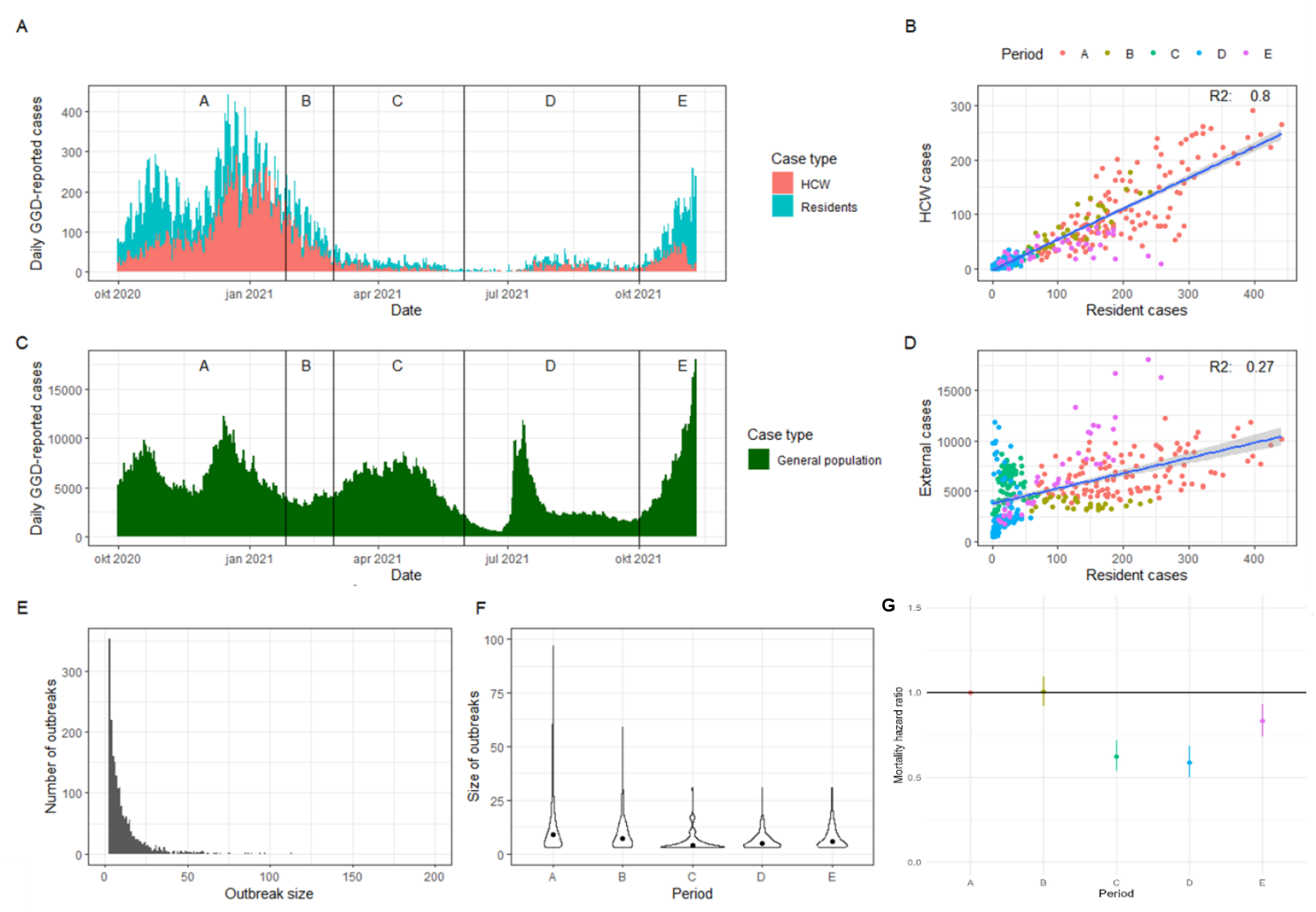
Descriptive results. **A) Epidemic curve based on reporting date in residents and HCW.** Vertical lines and number indicate study periods. **B) Correlation between daily number of reported resident and HCW cases**. The fitted linear regression model is shown with its corresponding R^2^ value. **C) Epidemic curve based on reporting date in the general population**. Vertical lines and number indicate study periods. **D) Correlation between daily number of reported resident and general population cases**. The fitted linear regression model is shown with its corresponding R^2^ value. **E) Histogram of outbreak sizes.** **F) Outbreak sizes per period**. Dots represent the median values. **G) Mortality hazard rate relative to period A**.

### Descriptive analyses: Outbreaks among residents in LTCFs

Resident cases for whom the LTCF identifier (68% of residents, n=25,086) was recorded could be grouped into outbreaks. Outbreaks were defined as three or more cases in the same LTCF notified within 14 days of each other. Of detected resident cases that could be matched to a specific LTCF, 88% were part of an outbreak. Overall, a total of 1984 outbreaks were detected in 1297 (73%) LTCFs, with 55% (n=980) of LTCFs reporting more than one outbreak during the study period (>2: 23%, >3: 6.7%). The maximum number of outbreaks reported in a single LTCF was six, which was reported by four LTCFs. Numbers of outbreaks per week across LTCFs varied over time, with most outbreaks per week occurring in period A (77 per week) and fewest in period C (7 per week). The median outbreak size was 7 (IQR 4-13) and lasted for a median of 14 days (IQR 7-23).

Across all outbreaks, 63.0% were considered large outbreaks (i.e., >5 cases), and 8.8% of outbreaks resulted in more than 30% of residents becoming infected (Figure 1E). There was no correlation between outbreak size and LTCF size (R^2^ = 0.02). Outbreak size varied significantly by period, with the largest median outbreak size observed in period A (median size = 9) and the lowest in period C (median size = 4) (Kruskal-Wallis rank sum test, p <0.001) (Figure 1F).

### Model-based analyses: Estimation of group-specific transmission rates

A subset of the individual-level data that contained bed capacity data (comprising 466 of the 1787 LTCFs; 26.1%) was used to estimate the contributions of different groups to the FOI in LTCF residents over time. For this subset, the median bed capacity of LTCFs was 71.5 (IQR 42 – 114). The total number of residents across these LTCFs was 39,150. LTCFs with available bed capacity were not equally distributed across the 24 GGD regions, with the number of LTCFs per region ranging from 4 to 38 (Pearson’s chi-squared test: p<0.001).

We used a Bayesian approach to estimate *β*_*i*_(*t*) for each group towards residents and quantify the contribution of each group to the overall FOI in residents. We compared model variants assuming different underlying distributions in the data and found a beta-binomial distribution to provide the best representation of the data (Supplementary figure 3 for a comparison of WAIC estimates). A model including a period term with only the significant interaction terms, which were the general population with period C and with period D, resulted in the lowest WAIC and was retained for further analysis (Supplementary figure 4).

The model presented a good fit to the observed data (Figure 2A, Supplementary figure 5) although the sizes of the largest outbreaks were underestimated. After correcting for the increased transmission potential of Alpha and Delta variants, we found that resident susceptibility was highest in period A and lowest in period C (62% reduction from period A), after which it increased to 57% of the initial susceptibility in period E (Figure 2D). Infectivity of HCWs was 32% lower compared to residents. Infectivity of the general population varied between periods, and was lowest in period C (59% lower than residents) (Figure 2E). We used the uncorrected estimates to calculate *β*_*i*_(*t*) from each group towards residents. From this we calculated resident-directed type-reproduction numbers (R) by multiplying *β*_*i*_(*t*) with the infectious period (5 days), (Supplementary table 2). The resident-to-resident R was 1.32 (95%CI 1.22-1.42) in period A, which declined to below 1 as COVID-19 vaccines were rolled out in period C (0.84 95%CI: 0.60-1.14), after which it increased again up to 1.50 (95%CI: 1.37-1.66) in period E (Figure 2B). A similar initial decrease was seen in resident-directed infections from HCWs with a 18% reduction between period A and B, and a 22% reduction between period B and C. The HCW-to-resident R was below 1 until period E. The largest observed difference was a 71% reduction between period B and C for the general population-to-resident R. Period C was the only period during which the resident-directed type-R from all three groups was below 1. Only during this period could the overall R of within-LTCF transmission be below 1.

**Figure 2:**
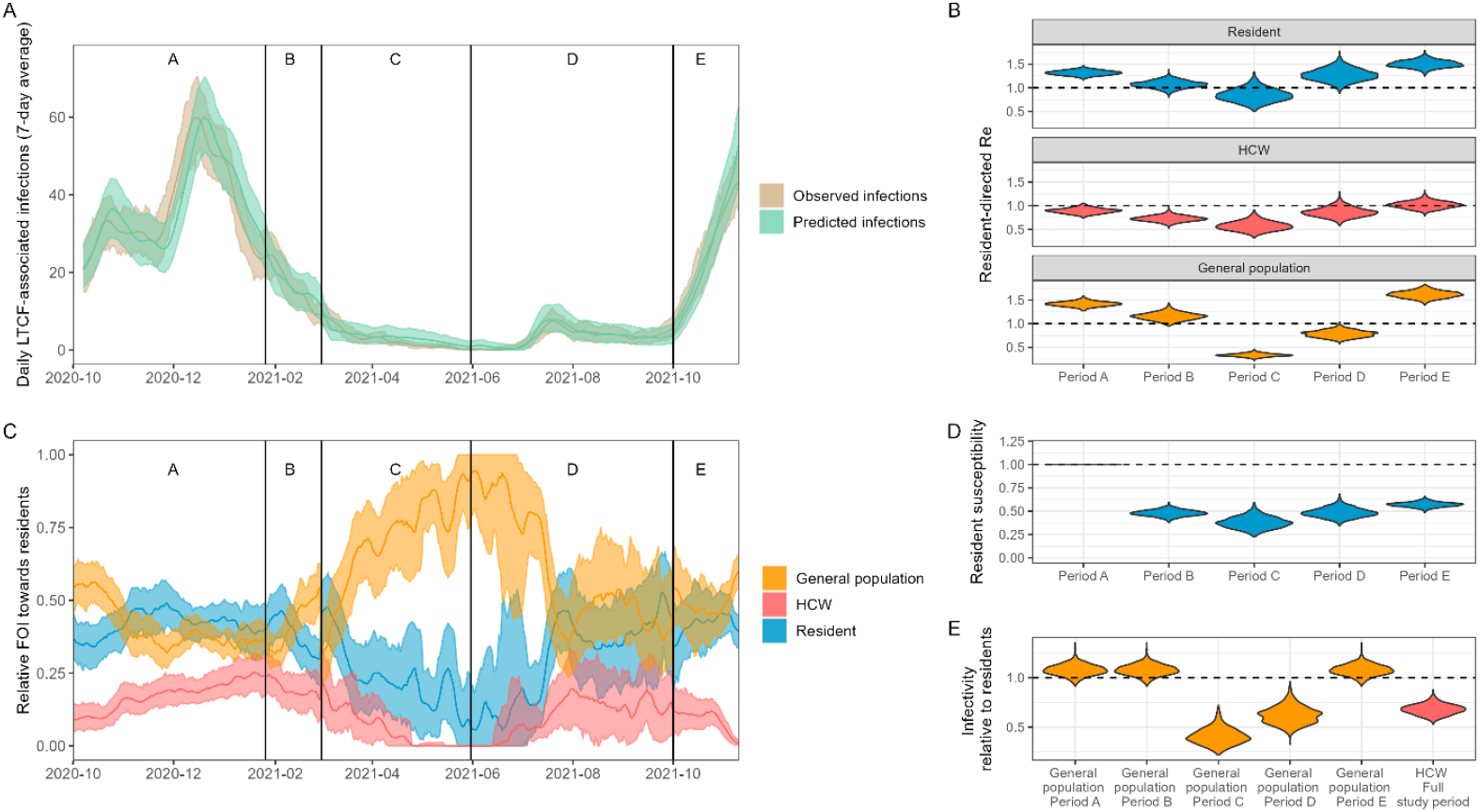
Model fits and outcomes for the best-fitting model. **A) Comparison between the daily number of observed infections (in the data) and predicted infections (from the model)**. Values of the 7-day average are shown. Dark lines indicate mean values. Shaded areas show 95% prediction intervals obtained by generating 1000 predictions by randomly sampling from the posterior distributions and augmented datasets. **B) Resident-directed R estimates for each LTCF group (resident/visitor/HCW)**. This was calculated as the product of the transmission rate parameter and the infectious period. **C) Daily relative force of infection from each LTCF group towards residents**. Dark lines indicate mean values. Shaded areas show 95% prediction intervals obtained by generating 1000 predictions by randomly sampling from the posterior distributions and datasets. **D) Relative susceptibility to infection in residents**. The horizontal line at y=1 indicates no difference compared to period A. **E) Relative infectivity compared to residents**. The horizontal line at y=1 indicates an infectivity equal to that of residents. Estimates for the infectivity of the general population differed in periods C and D compared to all other periods as these were significant interaction terms in the statistical model.

The overall daily FOI towards residents was highest in December 2020 (1.7*10-3, 95%CI: 1.5*10-3 – 1.9*10-3) and lowest in June 2021 (1.1*10-5 95%CI: 7.6*10-6 – 1.7*10-5) (Supplementary figure 9). We inferred that, in the pre-vaccination period (period A), 42.1% (95%CI 34.6% -48.8%) of the FOI experienced by residents could be attributed to fellow LTCF residents (Figure 2C). The contribution of the general population during this period was similar (40.8% 95%CI 35.1% - 46.9%). The contribution of residents to the FOI rapidly declined with the rollout of vaccination, with only 1 in 4 (23.7% 95%CI 11.6%-37.4%) infections in residents likely to result from another resident in period C.

HCW contributed least to the FOI across all periods, with a highest contribution of 19.7% (95%CI 13.2%-26.5%) in period B to the lowest in period C (5.8% 95%CI 1.7% - 11.2%). Like residents, the contribution of HCWs to the FOI declined with the rollout of vaccination, but for HCWs this did not return to pre-vaccination levels. The general population was the main source of infections in residents during period C (70.6%) and D (60.2%), when the absolute FOI was lowest and the uncertainty in estimates larger. These are the periods where residents and HCW had been vaccinated, but vaccination of the general population was still being rolled out.

When comparing the assumption of permanent, complete immunity in residents to assuming imperfect immunity in residents (default), we found that the values of the resident-directed type-R were on average 24% higher (supplementary table 3). Similarly, the mean FOI was on average 20% higher (supplementary figure 12). However, the relative FOI, relative resident susceptibility and relative infectivity of the other groups were robust to assumptions around immunity (supplementary figure 11). Assumptions regarding the extent of infection underreporting in the general population affected the R estimates related to this group, but none of the other outcome measures (supplementary table 4, supplementary figure 14 & 15).

## Discussion

We used a Bayesian GLM fitted to LTCF resident case data from autumn 2020 to autumn 2021 to quantify the contribution of residents, HCWs, and the general population to the force of infection in LTCF residents. We described characteristics of the COVID-19 epidemic in Dutch LTCF settings, with a focus on outbreaks among LTCF-associated cases. We observed the highest incidence rates as well as largest and longest outbreaks in the pre-vaccination period. Vaccination rollout was followed by a stark reduction in the average estimated infection susceptibility of residents. This reduction was robust to strong assumptions on the buildup of natural immunity, indicating that this reduction could not be explained by natural immunity alone. This protection decreased over time, which could partly be explained by increased transmission success of newly emerging Alpha and Delta variants. HCWs contributed least to the FOI towards residents over the study period, based on the information available regarding who is a LTCF HCW. Contributions from residents and the general population were similar in the pre-vaccination period, however, the general population formed the main source of infections in LTCF residents after vaccination was rolled out in LTCFs, which suggests a substantial reduction in within-LTCF transmission after vaccination. Within-LTCF resident-directed type-reproduction numbers were above 1 during most of the study period, meaning that outbreaks within LTCFs may occur. During the period shortly after vaccination, within-LTCF transmission may well have been self-limiting (i.e., R<1) provided that HCW-directed transmission was lower (or not much higher) than that directed towards residents.

The observed temporal trends in FOI contributions could be explained by several processes. During early spring 2021 (period B & C) Alpha became the dominant strain, followed by Delta in summer 2021 (period D & E) [31]. We detected no increase in transmission in LTCFs during the emergence of Alpha, which coincided with the rollout of vaccination in residents and HCWs. Estimates of temporal trends in residents’ susceptibility suggest that vaccine-induced immunity in the resident population was sufficient to counter the increased transmission success of the Alpha variant. Resident-directed R increased roughly six months after the first round of vaccination. This increase could be due to a combination of waning immunity [32]and the Delta variant becoming the dominant strain, which was associated with increased transmission success [33] such as through immune escape [34]. After correcting for the estimated transmission advantage of the Delta variant, estimates of residents’ susceptibility still increased towards the end of the study period, but remained low compared to those of the pre-vaccination period. These findings suggest that both waning immunity and the new variant played a role. Additionally, trends in transmission rate parameters could also be a consequence of seasonality [35], variation in case detection rates, and variation in contact between residents and all three groups due to additional control measures. Policies and the levels of adherence to control measures could have differed between LTCFs, which makes it difficult to study the effect of these interventions when looking at overall results. If detailed information on interventions and adherence to control measures would be available at LTCF or regional level, stratified analyses could shed light on their impact. Similar to observed trends in resident-directed R, mortality rates also started increasing towards the end of the study period, indicating that protection against mortality had also waned, possibly due to an increased mortality risk associated with the Delta variant, although its impact on mortality remains uncertain [36–38]. Mortality was recorded as all-cause mortality in the present study and likely suffered from underreporting due to the absence of a notification requirement. By estimating mortality rates relative to period A, we aimed to reduce the impact of this, although temporal changes in underreporting can bias our findings.

Our results indicate that a large proportion of infections in residents originated from the general population. Several other studies discussed the contribution of different groups to transmission in LTCFs, with significant variation in their findings. Genetic analyses of a large outbreak in a Dutch LTCF in the early phase of the pandemic indicated multiple introductions into the resident population, with a limited role for within-LTCF transmission [39]. Similarly, an outbreak in a Belgian nursing home originating from a single visitor showed a significantly increased infection risk in residents who attended the visitor’s event and limited onward transmission to those not attending [40]. Two model-based analyses from the United Kingdom [41, 42] also suggested the importance of external introductions, specifically by HCWs. Our results suggest a relatively small contribution of HCWs to the FOI towards residents. However, underreporting of infections in HCW is likely, e.g., due to misclassification of HCW status. An unknown part of the transmission from the general population should therefore probably be ascribed to HCWs or to other staff. Additionally, due to missing information about in which LTCF each HCW worked, we imputed this information when possible. This misclassification contributed to the uncertainty in HCW’s contribution to the FOI. Moreover, it has been shown that adherence to the reactive testing policy was higher for residents than for HCW, which indicates that asymptomatic infections in HCW were more likely to be missed [22]. It is also important to note that interventions such as masking were in place during the study period which could have contributed to the limited role of HCWs to transmission. Relative contributions to resident infections may have been different during the first wave, which was not in our study period, when fewer interventions were implemented.

Interventions are generally targeted towards reducing introductions into an LTCF, reducing transmission within an LTCF, or reducing the impact of infection. We showed that the estimated susceptibility of residents declined when vaccination in residents and HCWs was rolled out. This indicates that vaccination contributed to reducing introduction risk, limiting within-LTCF transmission, and reducing the impact of infections. This observed reduction in susceptibility, rather than in infectiousness, aligns with several studies showing that virus shedding was similar in vaccinated and unvaccinated individuals [43–45]. We only found statistical support for a reduction in infectiousness in the general population relative to residents in periods C and D (March-October 2021). While the general population tended to be more infectious than residents in periods A, B, and E, they were found to be less infectious than residents in periods C and D. The reason for this is unclear, but could be due to behavioral changes related to vaccination status or higher vaccine efficacy as the general population was generally younger [46, 47]. After the vaccination rollout in LTCFs, the general population was the dominant source of infections in residents and resident-directed Rs dropped below one. The variation in dominant sources of infection implies that the most effective control strategy for reducing infections in LTCFs varied over the course of the epidemic.

While we applied our model framework to LTCFs in the Netherlands, this approach could be easily adapted to other countries or similar settings where housing and healthcare are combined. Using a Bayesian approach combined with data augmentation allowed for a rigorous quantification of uncertainty and we showed a good model fit to the data. During a large-scale outbreak, dates of infection and case characteristics, such as an LTCF identifier, are often not observed or recorded. Data augmentation allows to explicitly account for these uncertainties in the analysis, making results robust to the impact of missing information. The availability of contact survey information for LTCF residents made it possible to adjust the prevalence in the general population to the prevalence in resident contacts (supplementary figure 2). While many other studies are limited by a lack of information on asymptomatic infections, the reactive testing policy in LTCFs resulted in less underreporting and allowed for detection of asymptomatic infections, although the adherence to this policy varied [22]. Asymptomatic individuals have been shown to contribute to transmission [48] but are challenging to detect, particularly in LTCFs, making this an important strength of this study. This was particularly important in later periods, when a larger share of infections lead to mild symptoms due to immune protection and are therefore more likely to be missed using symptom-based testing. Case affirmation rates among the general population are uncertain and may have varied due to variation in testing behavior over time and by age. To account for this, we assumed an underreporting proportion of 50% and restricted the study to the period where testing was available to everyone at sufficient capacity. Sensitivity analyses showed that deviations in this proportion affect R estimates for this population, but does not affect relative and absolute contributions to the resident-directed FOI from this group. A potential limitation is the incompleteness of data on LTCF capacity. However, there is no indication that this was related to regional or within-LTCF SARS-CoV-2 dynamics as this information predates the epidemic.

When designing interventions to reduce transmission it is important to characterize the epidemiological situation to inform the most effective strategy and avoid unnecessary harmful side-effects. We observed substantial variation over time in relative contributions of residents, HCWs, and the general population towards resident infections in LTCFs. This suggests that the effectiveness of potential interventions, which were focused on preventing introductions or within-LTCF transmission, varied over the course of the outbreak. This is valuable information that can be used for decision-making, such as the scheduling of vaccination rounds or the implementation of visitor restrictions to lower introduction risk. The ability to generate such insights relies heavily on robust surveillance systems, highlighting the importance of continued investments in these efforts. The application of advanced statistical techniques to routinely collected data opens opportunities to tailor interventions to the current epidemiological context and improve data-driven decision-making.

## Data Availability

Data hold by the RIVM can be requested by contacting. Other data mentioned in the manuscript needs to be requested by the named institutes, as these were provided under contract.

## Acknowledgments

We would like to thank Jantien Backer, Susan van den Hof, the RIVM modeling team, and the RIVM Covid-19 Epidemiology and Surveillance Team for their input to this study.

