## Supplementary material for "Contributions to the force of infection of SARS-CoV-2 in Dutch long-term care facilities"

### Supplement

**S1 Text. Methods**

**S2 Text. Results: Model selection**

**S3 Text. Results: Model fit and diagnostics**

**S4 Text. Results: Resident-directed type-R estimates**

**S5 Text. Results: Absolute FOI estimates**

**S6 Text. Results: Results: Resident susceptibility estimates without variant adjustment**

**S7 Text. Results: Sensitivity analyses immunity assumptions**

**S8 Text. Results: Sensitivity analyses underreporting infections in general population**

### 1. Methods

#### Data preparation

##### *Augmentation for LTCF-associated cases*

We augmented day of infection for resident and HCW cases to be able to calculate LTCF-level prevalence of infectious individuals. The day of infection was estimated based on available information on the timing of symptom onset or diagnostic test result.

*Day of infection.*

Following the observation process based of the LTCF reactive testing policy, we augmented the day of infection based on the day of symptom onset when an individual was the first reported case in an outbreak. We reconstructed the date of infection assuming the incubation period to be log-normally distributed with location parameter μ = 1.6, and scale parameter σ = 0.5 (median: 5 days) [1]. As the detection of a case led to the testing of all other residents, we used the day of positive test result for all subsequent people in same outbreak and for those with unknown day of symptoms onset. From the date of positive test result we reconstructed the date of infection using a normalized version of the probabilistic distribution of PCR positive test probability over time constructed by Hellewell et al. [2] (see *Supplementary Figure 1*). This distribution assumes that all exposures are detected between 0 and 30 days since exposure.

*Start and duration of infectiousness*.

We assumed that all cases started their infectious period three days after their day of infection [3] and that it lasted for five days, after which people start isolating. The duration of infectiousness was based on the incubation period, see above, and time between symptom onset and positive test in the Netherlands (2 days) [4].

*LTCF identifier.*
The LTCF identifier provides information about which LTCF someone lives or works. It was only available for a subset of residents (68%) and missing for HCWs. We imputed the LTCF identifier when missing for residents and HCWs by matching their cluster identifier to those from residents with recorded LTCF identifier. Cluster identifiers indicate which cases were linked together in space and time. Individuals across different LTCFs could be part of the same transmission cluster. When multiple LTCFs were associated with one cluster, we randomly selected, with equal weights, one of the LTCFs that were part of the cluster. This random selection was performed for each (augmented) dataset and thereby contributed to the variation between datasets.

##### *General population*

Missing information for the general population was imputed rather than augmented, because the number of cases in the general population was much higher than for LTCF-related cases thereby reducing the impact of individual variability on the outcomes. For those with recorded day of symptom onset, the date of infection was assumed to be five days earlier. This corresponds to the mean of the incubation period distribution described above. For those with recorded day of positive test result, the date of infection was assumed to be seven days earlier. This was based on the mean interval between symptom onset and positive test in the Netherlands (2 days) [4]. The start and duration of infectiousness was assumed equal to that of the LTCF-associated cases. We assumed an underreporting of cases in the general population of about 50%. This was estimated by comparing incidence estimates based on hospitalization data to the number of positive tests [5, 6]. Therefore, reported case numbers were multiplied by two to account for asymptomatic and unreported infections. We assessed the impact of this assumption in a sensitivity analysis.

We also required information on population sizes of each group to obtain daily prevalence estimates. Given the 1 to 0.98 resident to HCW ratio in Dutch LTCFs [7], we set the population size of residents and that of HCW equal to the LTCF capacity. To ensure the estimated prevalence in the general population better reflects the prevalence in those coming into contact with LTCF residents, we weighted age-specific prevalence estimates by the proportion of resident contacts within each age-group (see *Supplementary Table 1*).

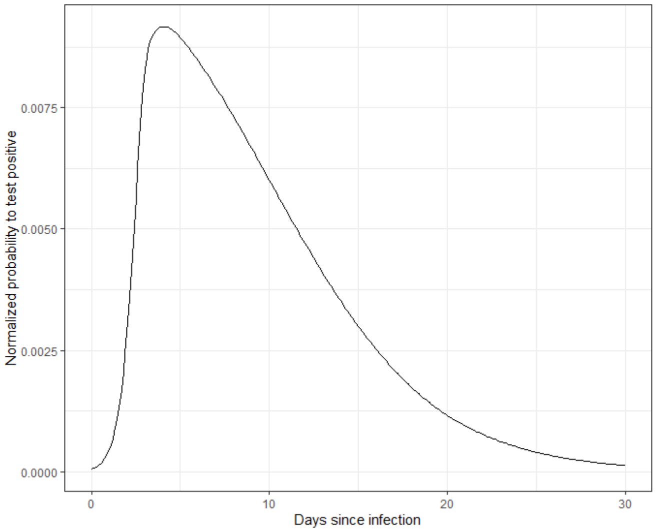


*Supplementary Figure 1: Distribution of time between infection and positive PCR test, adapted from Hellewell et al 2021.*

#### Age distribution of resident contacts

We analyzed data published in a contact study of people aged 70+ [8]. A total of 271 contacts were reported. The full age distribution is presented in Supplementary Figure *2* with the associated table for the age-distribution as used in the data from the general population in Supplementary Table 1.

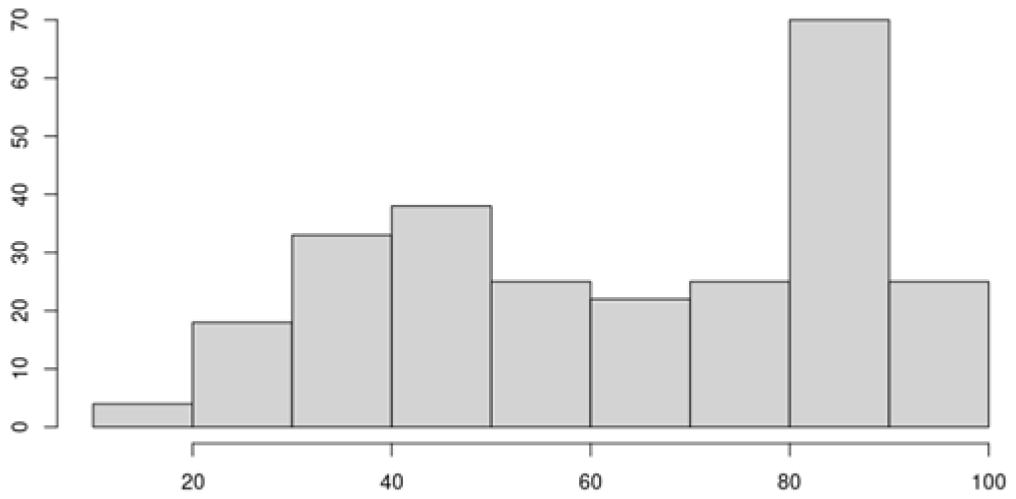


*Supplementary Figure 2: Age distribution of contacts of people aged 70+*

| Age in years | Proportion of resident contacts |
|---|---|
| **0-19** | 0.011 |
| **20-29** | 0.050 |
| **30-29** | 0.112 |
| **40-79** | 0.435 |
| **80+** | 0.392 |

*Supplementary Table 1: Contact matrix of LTCF resident contacts*

#### Estimation of time-varying transmission rate parameters

We considered that group-specific transmission rate parameters may have differed over time:

$$\beta_i(t) = exp^{c_0 + c_i x_1 + c_2 x_2 \ldots c_n x_n}. \qquad \text{(Equation 3)}$$

Here, $x_n$ and $c_n$ denote respectively the dummy variables and coefficients corresponding to each period $t$, with group 0 being the reference group whose effect is absorbed in the intercept $c_0$. Equation 3 implies that temporal changes in $\beta_i(t)$ were similar between groups. To relax this assumption, interaction terms between period and groups can be included.

#### Estimating relative susceptibility and infectivity

Relative susceptibility of residents and infectivity of HCW and the general population was estimated from the GLM coefficients. The susceptibility of residents (relative to period A) was obtained for each period by taking the exponent of the period coefficient ($c_p$). The infectivity of HCW was obtained by taking the exponent of the HCW coefficient ($c_{HCW}$). The same logic was applied to the general population, where the exponent of the general population coefficient ($c_{gen\ pop}$) was added to the exponent of the interaction term ($c_{gen\ pop\_period\_c}$, $c_{gen\ pop_{period\_d}}$) where applicable.

Adjustments for increased infectiousness of Alpha and Delta variants were made by dividing the resident susceptibility values by the increase in infectiousness (i.e., 1.7 in period B and C, 2 in period D and E).

#### 2. Results: Model selection

We compared WAIC values between models assuming different distributions in the data. Both the beta-binomial and the beta binomial zero-inflated distribution showed a large improvement compared to the binomial and binomial zero-inflated distribution, see Supplementary Figure *3*. As WAIC estimates using the beta-binomial and beta-binomial zero-inflated distributions were similar, the simplest model (beta-binomial distribution) was chosen.

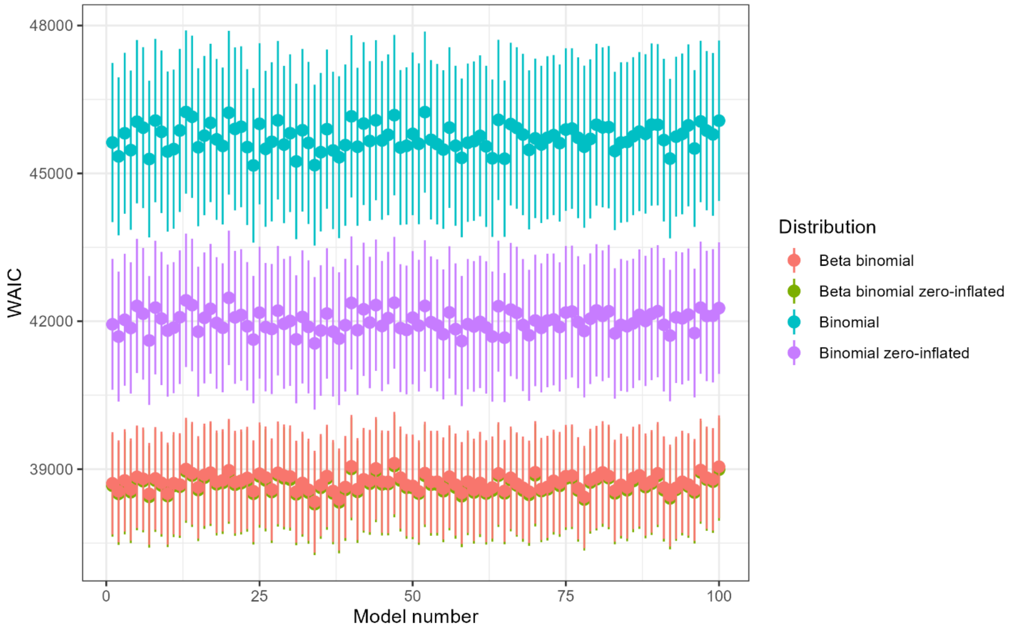


*Supplementary Figure 3: WAIC for each fitted model using different distributions. Vertical lines represent 95% confidence intervals*

Next, we compared WAIC values to assess the inclusion of period as an explanatory variable and its interaction with each group (i.e., HCW and general population). Models including a period term performed better than the null model, see Supplementary Figure 4. WAIC estimates of models with interaction terms performed better than those without, with the lowest WAIC values observed for the model only including the significant interaction terms. These were period C with the general population and period D with the general population. All further analyses were conducted on the model using the beta-binomial distribution including period as explanatory variable and these two interaction terms.

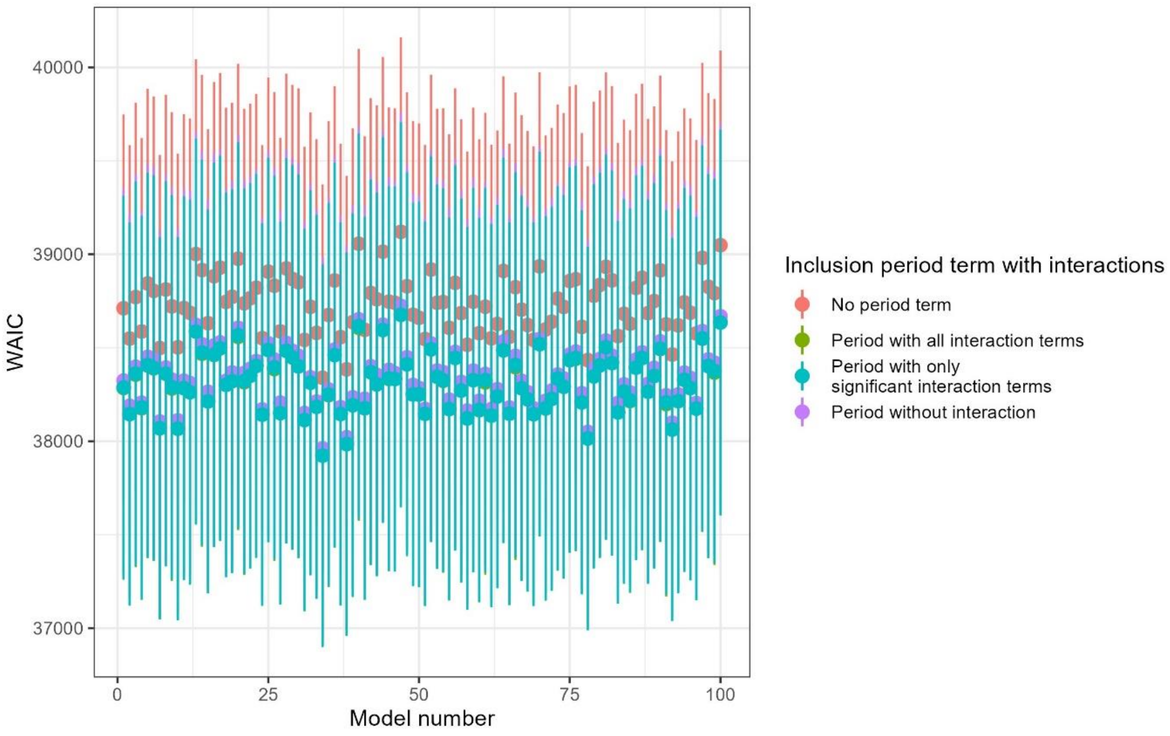


*Supplementary Figure 4: WAIC for each fitted model using different model formulations for the inclusion of period as explanatory variable with its interactions. Vertical lines represent 95% confidence intervals.*

#### 3. Results: Model fit and diagnostics

In addition to assessing model fit by comparing daily observed and predicted number of infections (main text figure 2A), we also compared the total observed and predicted number of infections per LTCF during the study period, see Supplementary Figure 55. Observations and predictions showed good agreement for small and medium number of infections, but sizes of the largest outbreaks were underestimated.

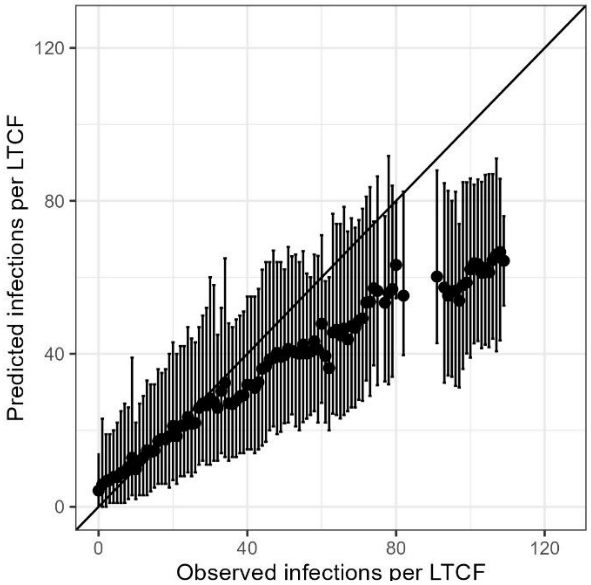


*Supplementary Figure 5: Comparison between number of observed infections per LTCF (in the data) and predicted infections per LTCF (from the model) over the full study period. The diagonal line indicates the points for which the observed value is equal to the predicted value. Dots indicate the mean predicted value. Errors bars represent 95% prediction intervals obtained by generating 1000 predictions by randomly sampling from the posterior distributions and augmented datasets.*

Traceplots, correlation plots and posterior density plots are shown below.

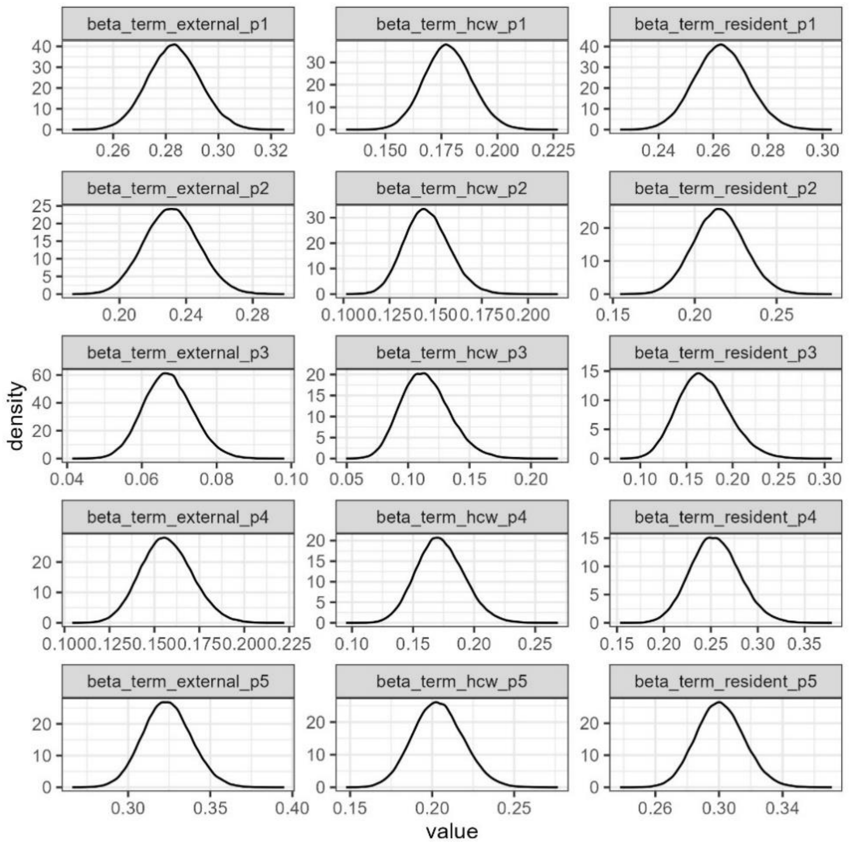


*Supplementary Figure 6: Posterior density plots of all transmission rate parameters*

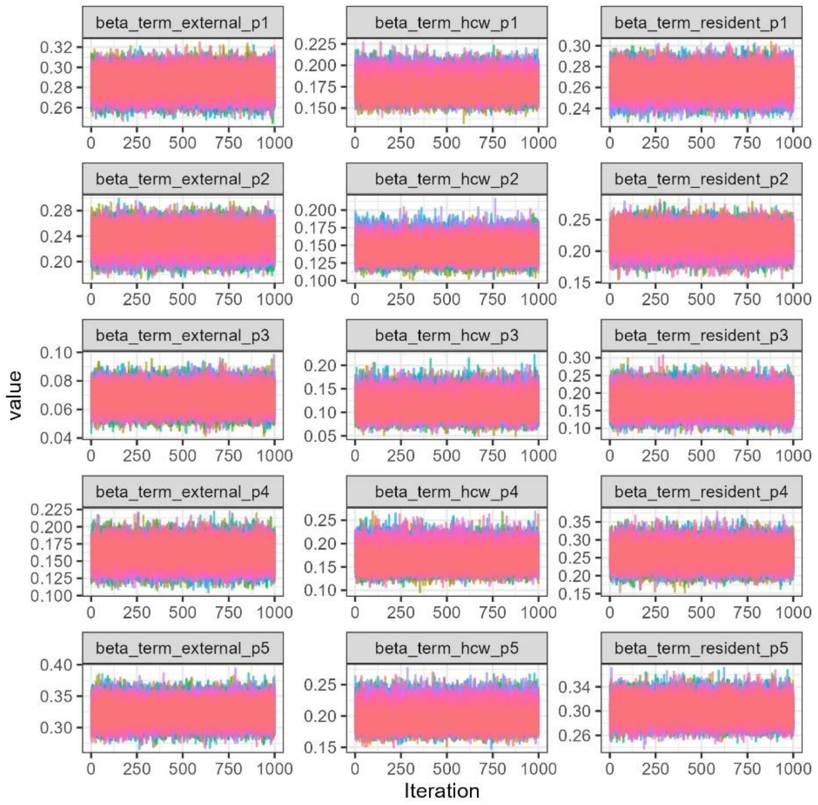


*Supplementary Figure 7: Traceplots of all transmission rate parameters*

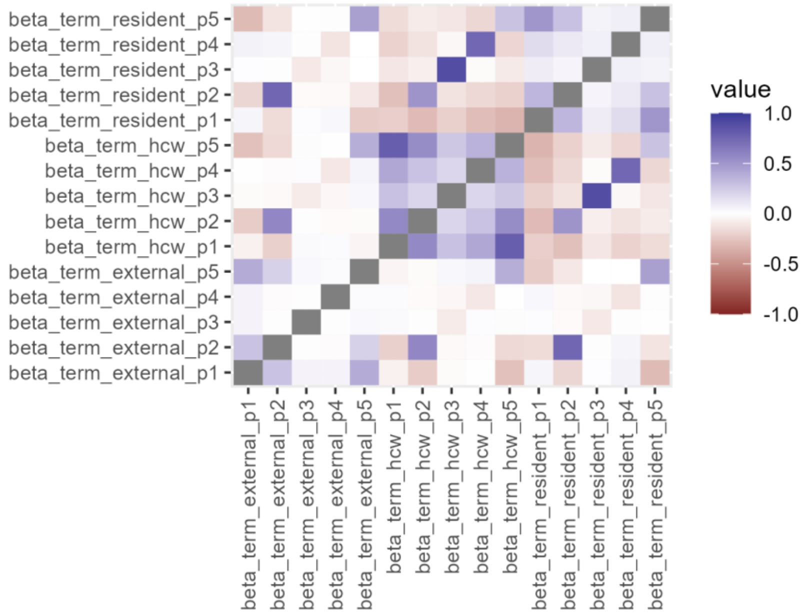


*Supplementary Figure 8: Correlation plot of transmission rate parameters*

#### 4. Results: Resident-directed type-R estimates

In addition to the visual presentation of the type-R values shown in main text figure 2B, the exact values are presented below for both the base model (not including variation over time) and the final (period) model.

*Supplementary Table 2: Estimates of resident-directed type-R for the base model and the period model, both using the beta binomial distribution. Mean WAIC is calculated as the mean across all augmented datasets (n=100). Phi is the overdispersion parameter.*

| | **Base model** | **Period model** | | | | |
|---|---|---|---|---|---|---|
| | | **Period A** | **Period B** | **Period C** | **Period D** | **Period E** |
| **Resident** | 1.34 (1.25-1.43) | 1.32 (1.22-1.42) | 1.08 (0.93-1.23) | 0.84 (0.60-1.14) | 1.27 (1.03-1.54) | 1.50 (1.37-1.66) |
| **HCW** | 0.88 (0.79-0.99) | 0.89 (0.79-1.00) | 0.73 (0.62-0.85) | 0.57 (0.40-0.78) | 0.86 (0.68-1.07) | 1.02 (0.88-1.18) |
| **General population** | 1.10 (1.04-1.17) | 1.42 (1.32-1.52) | 1.16 (1.00-1.32) | 0.34 (0.28-0.40) | 0.78 (0.65-0.93) | 1.62 (1.48-1.77) |
| **Mean WAIC** | 38,733 | 38,314 | | | | |
| **Phi** | 88 (82-96) | 89 (82-96) | | | | |

#### 5. Results: Absolute FOI estimates

In addition to the relative FOI estimates shown in main text figure 2C, the absolute values are shown below.

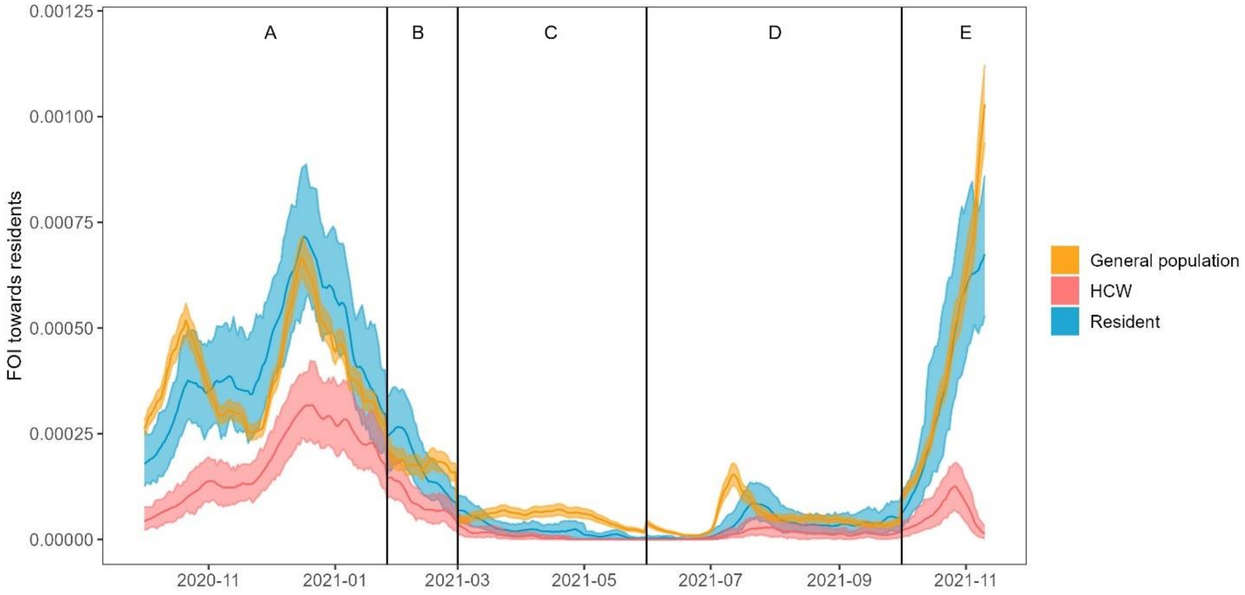


*Supplementary Figure 9: Daily force of infection from each LTCF group towards residents. Dark lines indicate mean values. Shaded areas show 95% prediction intervals obtained by generating 1000 predictions by randomly sampling from the posterior distributions and datasets.*

#### 6. Results: Resident susceptibility estimates without variant adjustment

Estimates for the resident susceptibility compared presented in the main text were adjusted for increased transmission success of newly emerging variants. Unadjusted susceptibility estimates are shown in Supplementary Figure 10.

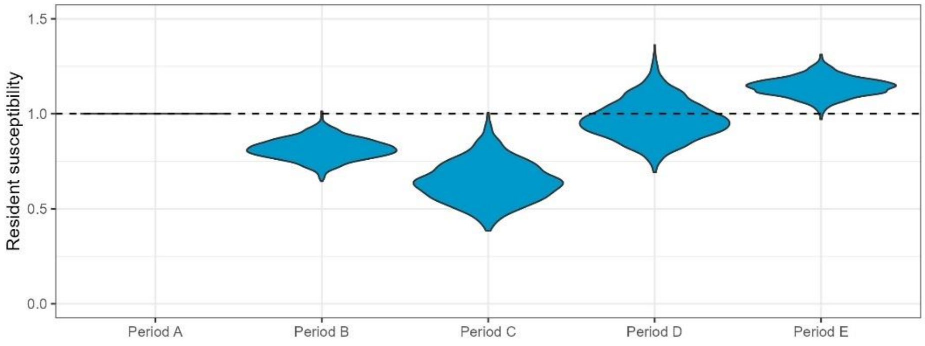


*Supplementary Figure 10: Relative susceptibility to infection in residents, unadjusted for increased transmissibility from newly emerging variants. The horizontal line at y=1 indicates no difference compared to period A.*

#### 7. Results: Sensitivity analyses immunity assumptions

We examined the impact of assumptions made about the size of the susceptible resident population. This is determined by model assumptions on the build-up of immunity in LTCF residents (i.e., whether complete immunity after infection is assumed), and the immune status of newly admitted residents (i.e., theoretically, following death of residents). The base scenario, as used in the main text, assumed imperfect immunity after infection, and that newly admitted residents were on average as susceptible as the resident population. In the sensitivity analysis, we modelled an extreme scenario where immunity after infection was perfect and lasting, and newly admitted residents were also immune. We assumed that (theoretical) new admissions happened on the same day following the death of residents.

This scenario differs in the number of susceptible people in each LTCF over time. For the baseline scenario, assuming imperfect immunity, the mean number of susceptible residents at an LTCF was 84, the median was 71. When assuming permanent, perfect immunity after infection with new residents also being immune, the mean was 75 and the median 62.

*Supplementary Table 3: Estimates of resident-directed type-R for the period model, using the beta binomial distribution. Immunity is assumed to be perfect and lasting, and new admissions comprise recovered residents. Phi is the overdispersion parameter.*

| | **Period model** | | | | |
|---|---|---|---|---|---|
| | **Period A** | **Period B** | **Period C** | **Period D** | **Period E** |
| **Resident** | 1.57 (1.46-1.69) | 1.44 (1.24-1.64) | 1.06 (0.75-1.43) | 1.57 (1.27-1.90) | 1.95 (1.77-2.15) |
| **HCW** | 1.09 (0.96-1.22) | 0.99 (0.84-1.16) | 0.73 (0.51-1.01) | 1.08 (0.86-1.34) | 1.35 (1.16-1.55) |
| **General population** | 1.56 (1.46-1.67) | 1.43 (1.23-1.63) | 0.40 (0.33-0.48) | 0.95 (0.79-1.13) | 1.94 (1.77-2.12) |
| **Phi** | | | 68 (63-74) | | |

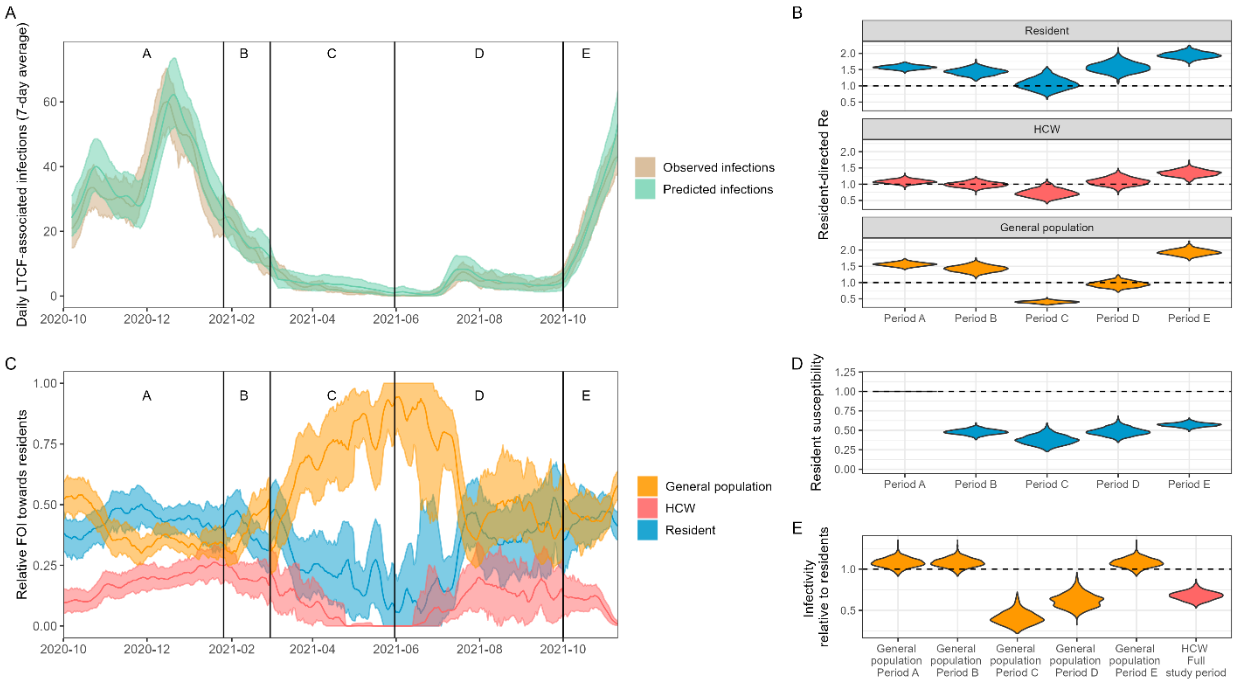


*Supplementary Figure 11: Model fits and outcomes for the alternative model assuming permanent and perfect immunity (sensitivity analysis).*

***A) Comparison between the daily number of observed infections (in the data) and predicted infections (from the model).*** *Values of the 7-day average are shown. Dark lines indicate mean values. Shaded areas show 95% prediction intervals obtained by generating 1000 predictions by randomly sampling from the posterior distributions and augmented datasets.*

***B) Resident-directed type-R estimates for each LTCF group (resident/visitor/HCW)****. This was calculated as the product of the transmission rate parameter and the infectious period.*

***C) Daily relative force of infection from each LTCF group towards residents****. Dark lines indicate mean values. Shaded areas show 95% prediction intervals obtained by generating 1000 predictions by randomly sampling from the posterior distributions and datasets.*

***D) Relative susceptibility to infection in residents.*** *The horizontal line at y=1 indicates no difference compared to period A.*

***E) Relative infectivity compared to residents.*** *The horizontal line at y=1 indicates an infectivity equal to that of residents. Estimates for the infectivity of the general population differed in periods C and D compared to all other periods as these were significant interaction terms in the statistical model.*

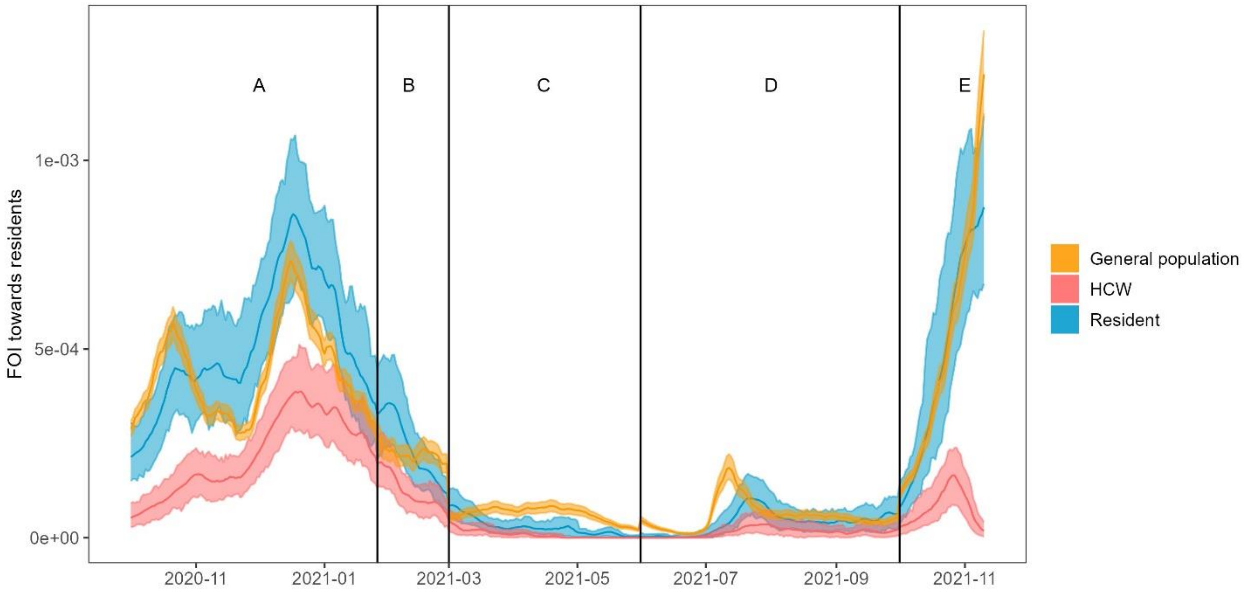


*Supplementary Figure 12: Daily force of infection from each LTCF group towards residents for the alternative model assuming permanent and perfect immunity (sensitivity analysis). Dark lines indicate mean values. Shaded areas show 95% prediction intervals obtained by generating 1000 predictions by randomly sampling from the posterior distributions and datasets.*

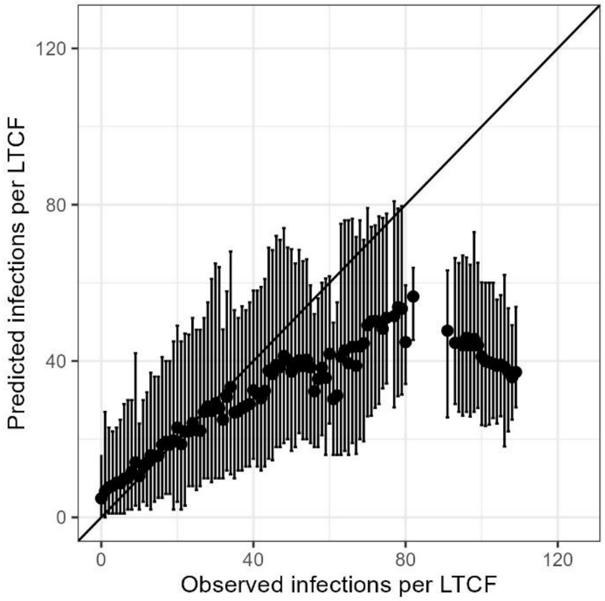


*Supplementary Figure 13: Comparison between number of observed infections per LTCF (in the data) and predicted infections per LTCF (from the model) over the full study period, for the alternative model assuming permanent and perfect immunity (sensitivity analysis). The diagonal line indicates the points for which the observed value is equal to the predicted value. Dots indicate the mean predicted value. Errors bars represent 95% prediction intervals obtained by generating 1000 predictions by randomly sampling from the posterior distributions and augmented datasets.*

#### 8. Results: Sensitivity analyses underreporting infections in general population

We examined the impact of assumptions made about the extent of underreporting of infections in the general population. In the main text, we assumed that 50% of infections went unreported and therefore multiplied the number of infections by 2. Here, we assumed that 75% of infections went undetected and we therefore multiplied the number of infections by 4.

Results showed that while estimates for the resident-directed effective reproduction number remain mostly unchanged for residents and HCW, the values have reduced by around 50% for the general population. All other outcome measures, including the contributions from the general population to the FOI experienced by residents were not affected by variation in underreporting rate.

*Supplementary Table 4: Estimates of resident-directed type-R for the period model, using the beta binomial distribution, assuming a 75% underreporting rate of infections in the general population. Phi is the overdispersion parameter.*

| | | | Period model | | |
|---|---|---|---|---|---|
| | **Period A** | **Period B** | **Period C** | **Period D** | **Period E** |
| **Resident** | 1.33 (1.22-1.44) | 1.08 (0.93-1.24) | 0.92 (0.64-1.29) | 1.30 (1.04-1.60) | 1.51 (1.36-1.67) |
| **HCW** | 0.85 (0.75-0.97) | 0.69 (0.58-0.82) | 0.59 (0.40-0.84) | 0.83 (0.65-1.05) | 0.97 (0.82-1.13) |
| **General population** | 0.73 (0.68-0.78) | 0.59 (0.51-0.68) | 0.17 (0.14-0.20) | 0.40 (0.33-0.47) | 0.83 (0.76-0.90) |
| **Phi** | | | 89 (82-96) | | |

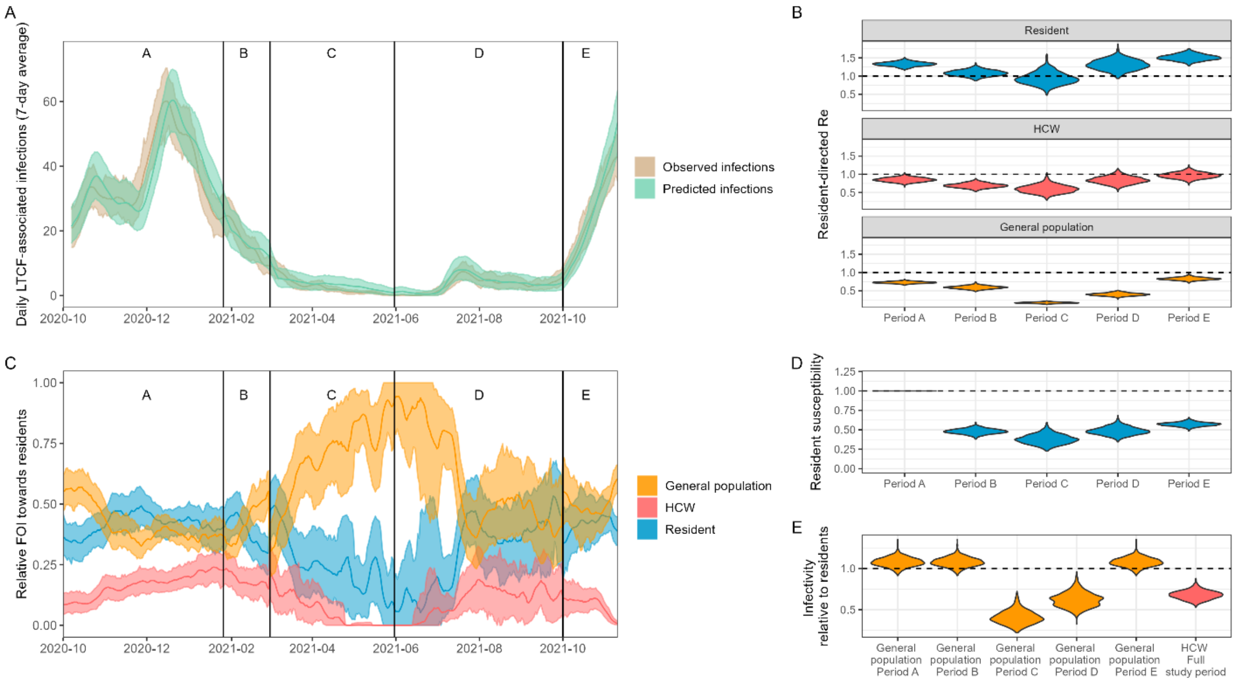


*Supplementary Figure 14: Model fits and outcomes for the alternative model assuming a 75% underreporting rate of infections in the general population.*

***A) Comparison between the daily number of observed infections (in the data) and predicted infections (from the model).*** *Values of the 7-day average are shown. Dark lines indicate mean values. Shaded areas show 95% prediction intervals obtained by generating 1000 predictions by randomly sampling from the posterior distributions and augmented datasets.*

***B) Resident-directed R estimates for each LTCF group (resident/visitor/HCW)****. This was calculated as the product of the transmission rate parameter and the infectious period.*

***C) Daily relative force of infection from each LTCF group towards residents****. Dark lines indicate mean values. Shaded areas show 95% prediction intervals obtained by generating 1000 predictions by randomly sampling from the posterior distributions and datasets.*

***D) Relative susceptibility to infection in residents.*** *The horizontal line at y=1 indicates no difference compared to period A.*

***E) Relative infectivity compared to residents.*** *The horizontal line at y=1 indicates an infectivity equal to that of residents. Estimates for the infectivity of the general population differed in periods C and D compared to all other periods as these were significant interaction terms in the statistical model.*

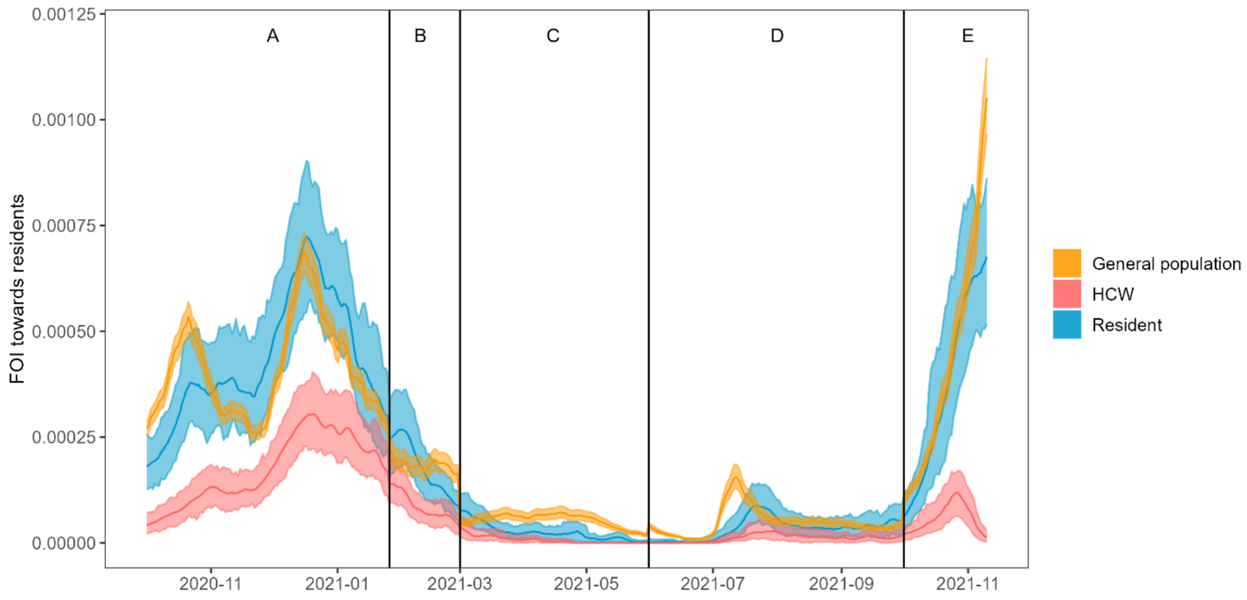


*Supplementary Figure 15: Daily force of infection from each LTCF group towards residents for the alternative model assuming a 75% underreporting rate of infections in the general population. Dark lines indicate mean values. Shaded areas show 95% prediction intervals obtained by generating 1000 predictions by randomly sampling from the posterior distributions and datasets.*
